# “We can’t just wish this thing away”: Caregiver perspectives on a child’s diagnosis of 3q29 deletion

**DOI:** 10.1101/2020.09.21.20198770

**Authors:** Megan R. Glassford, Ryan H. Purcell, Sarah Pass, Melissa M. Murphy, The Emory 3q29 Project, Gary J. Bassell, Jennifer G. Mulle

## Abstract

**Objective:** Genetic diagnoses are increasingly common in cases of intellectual disability and developmental delay. While ascertainment of a relatively common, well-studied variant may provide guidance related to treatments and developmental expectations, it is less clear how the diagnosis of a rare variant impacts caregivers, especially when the phenotype may include later onset manifestations such as psychosis. In the current study, we sought to identify caregiver concerns in the first qualitative study to assess the psychosocial impact of diagnosis on caregivers of individuals with 3q29 deletion syndrome (3q29Del), which is associated with a 40-fold increase in risk for psychosis.

**Method:** Participants were recruited from the national 3q29Del registry housed at Emory University (3q29deletion.org). Fifteen participants completed a semi-structured phone interview during which they were asked about their experiences before, during, and after their child received a diagnosis of 3q29Del. Interview responses were analyzed using the general inductive approach, and overarching themes were identified.

**Results:** We identified the following overarching themes: difficult “diagnostic odyssey,” mixed feelings about diagnosis, frustration with degree of uncertainty, and importance of resources. Importantly, our data suggest that future risk for psychosis is often not disclosed by medical professionals, consistent with the experience of individuals with 22q11.2 deletion syndrome.

**Conclusion:** These results highlight potential gaps in how caregivers are informed of risk for adult-onset conditions and point to key caregiver concerns for consideration in diagnosis of 3q29Del.

## INTRODUCTION

Early and accurate diagnosis of a child with developmental delay is important to caregivers and health care providers. Technological advances have dramatically improved the molecular diagnostic yield as compared to karyotype analysis and have thereby also changed the diagnostic experience for caregivers and families.^1^ However, today a molecular (genetic) diagnosis can elicit more questions than answers. While a molecular diagnosis can provide a measure of etiologic validation, many genetic variants linked to neurodevelopmental delay are rare and poorly understood and therefore may have little current prognostic utility.

One such rare neurodevelopmental disorder is 3q29 Deletion Syndrome (3q29Del, MIM #609425), a recurrent hemizygous deletion of 1.6Mb and 21 protein-coding genes, which has an estimated population frequency of 1 in 30-40,000 and typically arises *de novo*.^2-4^ 3q29Del was first described in six patients with mild to moderate intellectual disabilities.^2^ The phenotypic spectrum has expanded as more affected individuals have been identified but is still under investigation.^5^ In addition to intellectual disabilities, some of the most common clinical features include developmental and speech delay, low birth weight, failure to thrive, and short stature.^2- 4,6^ Like other several copy number variants (CNVs) such as 22q11.2 deletion, 3q29Del has also been shown to be associated with extremely high risk for neurodevelopmental and psychiatric disorders including schizophrenia (OR > 40),^7-9^ autism spectrum disorder (ASD),^10-13^ bipolar disorder,^10,14^ obsessive compulsive disorder,^11^ and clinical depression.^15,16^ In the 3q29Del patient registry housed at Emory University (3q29.org), 28% of study subjects report diagnosis of at least one psychiatric condition.^6^ A more recent analysis found that of 93 study subjects with 3q29 deletion, 59% reported global developmental delay/mental retardation, 28% report generalized anxiety disorder, and 29% reported autism spectrum disorder.^12^ Importantly, there is not a known hallmark characteristic of 3q29Del.^4,17^ These data highlight the high risk that 3q29Del confers for a broad spectrum of neurodevelopmental and neuropsychiatric illness. The current relative paucity of phenotypic and prognostic data for 3q29Del in contrast to the stark risks for neuropsychiatric disease may unexpectedly alter the diagnostic experience for caregivers.

Previous research has shown that caregivers value finding a specific, medically recognized diagnosis for their child, but long diagnostic odysseys can have a negative impact on caregivers and patient.^18-20^ An early, official diagnosis can facilitate access to services, which may improve health outcomes, and patients and families may benefit from support services.^19,21^ 3q29Del-associated intellectual disability is typically mild to moderate and unlike other genetic syndromes, facial dysmorphology is subtle and does not serve to distinguish the presence of the deletion. Indeed, many of the manifestations are behavioral. Anecdotally, caregivers of these children report facing blame, recrimination, and accusations that they are “enabling” their child’s behavior. These judgements come from strangers, school professionals, and even well-meaning family members. In this context, the importance of a diagnosis takes on a new significance for caregivers. We therefore sought to identify these issues using a qualitative study design.

The present study seeks to understand 3q29Del caregiver perspectives on the impact of diagnosis, which will provide insight into caregiver experiences and inform resource needs and targets for healthcare providers (HCPs) who work with these populations.

## METHODS

### Participants

Study subjects were recruited from the 3q29Del registry (Emory University, 3q29.org) and a 3q29 caregiver-run Facebook page. The registry was launched in 2013 and has ascertained data on the largest known sample of individuals with 3q29Del.^6,12^ Recruitment was accomplished with a personalized email to registry participants describing the study and requesting follow-up contact by willing participants. Additionally, study details and similar contact instruction were posted on the 3q29Del Facebook page. To be eligible for inclusion, study subjects had to be a primary caregiver of an individual with 3q29Del and a native English speaker. Sixteen individuals expressed interest but one person was excluded as a non-native English speaker. The remaining 15 individuals consented and participated in the study.

### Procedures

Interested prospective participants were sent a follow-up email to schedule the in-depth, individual interview with interview questions included. Participants provided informed oral consent prior to the interview. Semi-structured interviews were completed by phone and audio was recorded. This study was approved by the Emory University Institutional Review Board (IRB00064133).

### Instrumentation

Interview guide questions were open-ended and exploratory in nature (Table S1) and addressed feelings and experiences as a caregiver of a child with 3q29Del. Questions were broken down into three sections: pre-diagnosis, time of diagnosis, and post-diagnosis. Probes and follow-up questions were used as necessary to generate rich data. The interview guide also included demographic questions and additional demographic information was obtained from participant profiles in the 3q29 deletion registry. Interviews ranged from 20-60 minutes and participants were compensated with a $25.00 gift card.

### Data Analysis

We used general inductive analytical approach.^22^ Interviews were transcribed verbatim and uploaded into MAXQDA (VERBI Software), a qualitative analysis software program.^23^ Deductive codes were identified prior to analysis based on the literature and experience with registry participants. All interviews were then analyzed line by line and inductive codes were identified and assigned to segments of text through the reading of the raw data. New codes and subcodes were identified until data saturation.

All interviews were independently coded by two experienced qualitative researchers. Any differences in coding were discussed and reconciled between the two coders. Codes were then collapsed into overarching themes.

## RESULTS

The majority of the 15 participants were Caucasian (86.7%) and married (86.7%), and all were female and highly educated (Table I). The median age of participants was 39 years, and the median current age of the affected child was 7 years. The median age of diagnosis for the affected child was 3 years. High inter-coder reliability was achieved (Cohen’s kappa^24,25^ statistic = 0.80). Three deductive codes were identified prior to the coding process, and an additional thirty inductive codes were identified throughout the coding process for a total of thirty-six codes. We identified four overarching themes: 1) difficult diagnostic odyssey, 2) mixed feelings about the diagnosis, 3) frustration with degree of uncertainty, and 4) importance of resources, each of which are described in detail below. Salient quotes illustrating the themes are provided in Table II.

**Table I.** Demographics.

|  | **Categories** | **Frequency (%)**<br>**N=15** | **Median**<br>**(Interquartile Range)** |
| --- | --- | --- | --- |
| **Caregiver Ethnicity** | Caucasian<br>Caucasian/South African<br>Native American | 13 (86.7%)<br>1 (6.7%)<br>1 (6.7%) | N/A |
| **Caregiver Gender** | Female<br>Male | 15 (100%)<br>0 (0%) | N/A |
| **Caregiver Highest Level of Education** | Undergraduate Training<br>Graduate Training<br>Doctorate Training | 8 (53.3%)<br>6 (40.0%)<br>1 (6.7%) | N/A |
| **Caregiver Marital Status** | Single<br>Married<br>Divorced | 1 (6.7%)<br>13 (86.7%)<br>1 (6.7%) | N/A |
| **Child's Age at Diagnosis** | N/A | N/A | 3 years<br>(5 years) |
| **Age of Affected Child** | 0-5 Years<br>6-10 Years<br>11-15 Years<br>16-20 Years | 6 (40.0%)<br>4 (26.7%)<br>4 (26.7%)<br>1 (6.7%) | 7 years<br>(10 years) |
| **Caregiver's Age** | 20-25 Years<br>26-30 Years<br>31-35 Years<br>36-40 Years<br>41-45 Years<br>46-50 Years | 0 (0%)<br>1 (6.7%)<br>6 (40.05)<br>2 (13.3%)<br>2 (13.3%)<br>4 (26.7%) | 37 years<br>(14 years) |
| **Caregiver's Age at Diagnosis** | 20-25 Years<br>26-30 Years<br>31-35 Years<br>36-40 Years<br>41-45 Years<br>46-50 Years | 1 (6.7%)<br>3 (20.0%)<br>5 (33.3%)<br>2 (13.3%)<br>4 (26.7%)<br>0 (0%) | 35 years<br>(12 years) |
| **Caregiver's Total Number of Children** | 1<br>2<br>3<br>4<br>5 | 1 (6.7%)<br>5 (33.3%)<br>3 (20.0%)<br>5 (33.3%)<br>1 (6.7%) | 3 children<br>(2 children) |

**Table II.** Overarching Themes.

| Description of Theme | Example Quotes |
| --- | --- |
| **Difficult diagnostic odyssey**<br>Related to degree and specificity of symptoms as well as available technology | *“I mean I noticed that there were some significant delays...and we actually had gone to his doctor several times, and I had... asked if those things were...atypical or not and the doctor, his pediatrician, said ‘oh no, he’s fine.’” – Int. 3*<br>*“The frustrating part for the first couple years was knowing that something wasn’t right but not knowing what it was and not knowing what to do to help or not knowing what to do to fix it.” – Int. 2* |
| **Mixed feelings about diagnosis**<br>Validation, relief, frustration, despair | *“ when we got the diagnosis, it was more of a relief because I was like I knew it...I mean it was upsetting at the time because it’s like, okay, now he really does have something and we can’t just wish this thing away...but at the same time, we were just like, I knew it, I told y’all there was something comprehensive going on with him.” – Int. 3*<br>*“I was like another thing? He already has a massive heart defect. Now, we have to do this...so, I had those feelings, but then, on the flip side, I felt like I also had an answer to why he was so delayed. And, so, on that side, I felt relieved.” – Int. 11* |
| **Frustration with degree of uncertainty**<br>Variable phenotypic spectrum, risk for psychosis, level of independence, long-term prognosis | *“It’s somewhat discouraging because there’s not enough known to know exactly what he’s capable of.” – Int.2*<br>*“We can’t tell you if he’s going to be functional. We can’t tell you if he’s gonna talk...We can’t tell you anything. That’s...always, for me, been the hardest.” –Int. 9*<br>*“people don’t even know about what to expect...with this syndrome, and I know it’s very varied, so like, I just feel like I don’t even know...should I be saving up for lifelong care or college?” – Int. 12* |
| **Importance of resources**<br>Knowledgeable and motivated HCPs, support of other affected families, access to therapeutic resources | *“I think it’s always nice to know that you’re not the only person going through the things.” – Int. 6 – Int. 6*<br>*“As soon as he was born, my [other] kids went from having one parent to having no parents because he took every ounce of energy and everything I have.” – Int. 2*<br>*“our insurance had basically said that...we know that there’s something wrong with him but there’s not something wrong enough that we’re going to pay for it.” – Int. 9* |

### Difficult Diagnostic Odyssey

The theme *difficult diagnostic odyssey* encompasses caregivers’ emotional and financial experiences related to searching for a diagnosis for their child. Several caregivers noticed that something was different about their child at birth or shortly thereafter. Concerns often included developmental delays, feeding problems, and/or differences in social behaviors. However, caregivers reported that their concerns were often not taken seriously by HCPs. Additional reasons for delayed diagnosis included the child exhibiting mild symptoms (such as feeding difficulties) or symptoms that lacked specificity (such as global developmental delay). Caregivers expressed that this forced them to be strong advocates for their child. As one caregiver put it, “it was lots of trial and error. It was lots of different doctors. It was lots of medications.” Additionally, this led to financial hardships as insurers often refused to cover the costs of these necessary interventions and therapies due to the lack of a supporting diagnosis.

Due to these delays, some caregivers felt that finding the exact diagnosis was secondary to improving their child’s level of health and ability to function. Alternatively, some caregivers explained that they were in denial and assumed the child would eventually “grow out of it.”

Caregivers also mentioned the state of genetic testing technology at the time diagnostic tests were ordered. In some cases, a genetic condition had been suspected early on by HCPs and genetic testing was ordered for specific conditions, but the results were negative. Thus, some doctors were hesitant to order additional genetic testing. Insurance companies also refused to cover additional genetic testing for some children despite technological advances. Caregiver reactions to previously negative genetic testing results varied. Some still felt sure that there was an underlying genetic condition and became frustrated and confused with HCPs delivering multiple diagnoses to describe multiple individual symptoms and features rather than a unitary diagnosis that addressed the overall picture, and possibly, the underlying cause.

### Mixed Feelings about Diagnosis

The theme *mixed feelings about diagnosis* illustrates caregivers’ reactions and, at times, contradictory feelings about the child’s diagnosis. Overall, most (9/15) caregivers felt a mix of both positive and negative feelings after receiving the diagnosis. For those who had been sure their child had an underlying genetic condition, they initially felt validation and relief. For some, having a diagnosis resulted in caregivers feeling they had a greater understanding of their child’s condition, including what expectations they should have moving forward. Another described benefit of diagnosis came in determining what types of treatments and interventions were most appropriate for their child. Importantly, the diagnosis was helpful in accessing and obtaining insurance coverage for treatments and interventions, which had been a significant financial burden before having a diagnosis.

We observed some slight differences in caregiver feelings about their child’s diagnosis, which was related to their level of difficulty with finding a diagnosis. Overwhelmingly negative feelings about the diagnosis, including fear and despair, were more likely to be expressed by caregivers who received a diagnosis for their child shortly after they first expressed concerns. In contrast, overwhelmingly positive feelings, including validation and relief, were expressed by two caregivers, one who endured a difficult diagnostic odyssey and one who did not.

Some caregivers viewed this diagnosis specifically as being hopeful because they felt 3q29Del is much less severe than many other possible genetic diagnoses. In contrast, others felt of despair and hopelessness after learning the genetic diagnosis. Additionally, 13% (2/15) openly expressed that either their partner or themselves experienced feelings of parental guilt after learning about the diagnosis. In some cases, the value of having a diagnosis was questioned because so little information about 3q29Del is available.

### Frustration with Degree of Uncertainty

The theme *frustration with degree of uncertainty* highlights the lack of HCP awareness and available information about the phenotypic spectrum and long-term prognosis of 3q29Del. When caregivers first learned about their child’s diagnosis, many expressed that they were frustrated that their HCPs were not able to answer questions or provide more information about what this diagnosis means for their child. Furthermore, when discussing the diagnosis with other HCPs, caregivers found themselves taking on the role of educator because most HCPs were unfamiliar with 3q29Del and the associated health concerns. As one caregiver said, “The diagnosis came back that he has 3q29 microdeletion…it was the first one the doctor had ever seen.” The lack of data and scientific literature was also disappointing for caregivers who wanted to do their own research after diagnosis. Following the diagnosis, one parent remembered “that night like I think I came home and read everything that was available on it. And, so, by the time I talked to the genetic counselor, I knew more than she did.”

The lack of information made some caregivers question the value of this diagnosis. “I don’t know whether getting a name when the condition is still unknown, especially back then, with such little information on it. I’m not sure that that really helped at the time.” Caregivers expressed that the range and severity of possible symptoms is so broad that they felt like they still did not have an answer for what to expect. One participant wondered “Should I be saving up for lifelong care or college?” On the other hand, some caregivers found a sense of hope after learning about the variable phenotypic spectrum. They explained that they felt “lucky” because that their child seemed to be on the mild end of the spectrum.

One significant concern that was described by 67% (10/15) of caregivers was the association with psychiatric illnesses. For children who have already manifested psychiatric symptoms, these were described as one of the most troubling aspects of the syndrome. For children who have not yet shown any signs or symptoms of a psychiatric illness, caregivers expressed that they often worry about if and when the child will develop these symptoms.

### Importance of Resources

*Importance of resources* as a theme brings together the educational, therapeutic, and support resources that can be helpful for affected individuals and family members. Most caregivers (93%; 14/15) were provided with some type of educational resource to take home with them at the time of diagnosis. Most often, this was a brochure on 3q29Del from the Unique support group.^26^ Although 73% (11/15) of caregivers stated that they understood the information provided at the time of diagnosis, some felt overwhelmed and confused by the information. Others felt the diagnosis was not explained adequately or that not enough literature was provided.

The stress described by caregivers highlighted the need for resources. Causes of parental stress included: enduring a long diagnostic odyssey, providing interventions for their child, physical and emotional demands of caring for a child with special needs, and the impact of their child’s health problems on daily life activities and plans for the future. The risk for other and/or future children to be affected was a concern and source of stress mentioned by 40% (6/15) of caregivers. The effect on siblings of this diagnosis was mentioned by some caregivers. These concerns were related to the potential impact on siblings’ emotional health and the potential need for siblings to become the caregivers in the future.

Caregivers listed helpful resources such as a knowledgeable, multidisciplinary healthcare team in place, the Emory 3q29 patient registry, 3q29 Facebook group, and other support groups. Many caregivers explained that the private Facebook group was the most beneficial resource highlighting the benefits of peer resources but also perhaps underscoring the lack of available resources from HCPs. Eight out of fifteen caregivers (53%) also expressed a desire to help other affected children and families by helping to increase awareness of 3q29Del by participating in the patient registry and other research studies.

## DISCUSSION

These findings indicate that the molecular diagnosis of a rare variant associated with neuropsychiatric disorders presents multiple challenges for caregivers and families. In the case of 3q29Del – which may be relevant to a host of other rare genetic syndromes – little is known at this time about the syndromic attributes or patient prognosis, no treatments options are known to be effective, and many providers are unfamiliar with the condition. As a result, the ascertainment of a genetic variant does not necessarily provide a satisfactory conclusion to the diagnostic odyssey for caregivers but rather can give way to new journey from pediatric to adult neuropsychiatric specialists.

Many similarities were noted between the findings of this study and similar studies in other patient populations particularly under the theme of difficult diagnostic odyssey. For example, in the time period prior to diagnosis, caregivers described barriers to reaching a diagnosis of 3q29Del, including parental concerns not being taken seriously, a lack of awareness among HCPs, a lack of specificity of symptoms, and limitations of genetic testing technology. Similarly, caregivers of children with 22q11.2 deletion syndrome (22q11DS) also reported that their concerns were not taken seriously by HCPs,^27^ and lack of awareness was identified as a barrier by caregivers of children with fragile X syndrome.^28^ Additionally, caregivers of children with 3q29Del explained that if the child was born prematurely or with a birth defect, such as a heart defect, then developmental delays and other parental concerns were dismissed and attributed to these prior conditions. In 3q29Del and in other genetic syndrome patient populations, including fragile X syndrome, it is common for the child to receive multiple diagnoses prior to receiving the genetic diagnosis.^28^ Prior diagnoses typically described manifestations of the condition rather than the underlying genetic etiology and often included ASD, global developmental delay, and failure to thrive. Limitations of genetic testing technology also played a role in delayed diagnosis in some cases. Many individuals in this study had previously received a negative result to a genetic test, often a reflection of the technical limitations of the test, or its specificity toward a more common syndrome such as 22q11DS or fragile X syndrome. These barriers and delays in diagnosis made it necessary for parents to advocate for their child to receive referrals to specialties, therapies, and genetic testing, which has also been described in other similar studies.^28^

At the time of diagnosis, caregivers described mixed feelings including validation, relief, frustration, and despair. These feelings are also consistent with what has been described in similar studies.^27-29^ Caregivers of children with fragile X syndrome have reportedly felt both relief and devastation.^28^ Similarly, a study of 22q11DS described “ambivalence between relief and sorrow” about the diagnosis and that parents who had been searching for a diagnosis for several years felt relief and validation whereas parents whose child received a diagnosis at an early age felt more overwhelmed and found it difficult to accept that their child had an underlying condition.^27^

Frustration with the uncertainty of this diagnosis was an overarching theme in this study, which describes caregivers’ feelings at the time of diagnosis as well as post-diagnosis. This uncertainty stems from the variable phenotypic spectrum of 3q29Del and the lack of understanding among HCPs. Although some caregivers were thankful for any amount of information, some questioned the value of this diagnosis in line with a previous report.^29^ These sentiments highlight the paradox of ascertaining a genetic disorder before the syndrome is well-characterized.

The association with psychiatric illnesses, and schizophrenia in particular, is another aspect of uncertainty and point of concern for caregivers. However, this association was not always disclosed at the time of diagnosis. Similarly, a study of 22q11DS found that 61.5% of caregivers were not informed of this association.^30^ Consistent with our findings, the possibility of their child developing a psychiatric illness was a great source of anxiety for most of caretakers. Although this association was a major concern among caregivers in both studies, there was a difference in the root of these concerns. Hercher et al. (2008) found that possible stigmatization was a main contributing factor to this heightened anxiety. In our study, caregivers were more concerned with how a psychiatric illness would impact quality of life and level of independence. Overall, these findings highlight the importance of disclosing the risk for psychiatric manifestations and providing appropriate educational and support resources at the time of diagnosis in order to help caregivers prepare for and cope with this possibility.

Overall, the lack of information creates a “fear of the unknown,” which pertains to expectations for what the future holds for their child as far as their health, psychological state, and level of functioning and independence. Thus, caregivers worry about what they should be doing to prepare for the future, a future that may end up being much different than they originally planned. In addition, some HCPs were perceived by caregivers as being uninterested in learning more about 3q29Del, which they felt interfered with their child’s health care. Stemming from this, caregivers described a lack of identity and community for families affected by 3q29Del compared to more well-known genetic conditions.

Caregivers for children with special needs experience significant stress and this strain can also impact other aspects of family structure.^27^ Moving forward after learning of their child’s diagnosis, caregivers felt that the most helpful resources were those that allowed them to ask questions and share stories with other affected families, such as the Facebook group. These types of support resources also help caregivers to know they are not dealing with this diagnosis alone. Thus, while it is important to provide educational and therapeutic resources, support for the affected child, caregiver, and family members, especially siblings should be considered.^27,29^

### Study Limitations

Based on the qualitative nature of the study, our findings cannot be considered to represent all caregivers of children with 3q29Del. The possibility of participation bias and/or ascertainment bias is a limitation of this study. As all participants were women, these results are likely only representative of maternal caregiver experiences. It is possible that the results could have differed if men had participated in the study. Caregivers with strongly positive or strongly negative experiences may be more likely to participate in the registry, Facebook group, and/or this study. Individuals enrolled in the registry and Facebook group may also have children who are more severely affected. Thus, this study may over-estimate caregiver concerns. However, if changes are adopted to serve the most severely affected families, it is likely these changes will be beneficial in improving the experiences of families with less severely affected children as well. Furthermore, the registry and Facebook group are web-based and therefore may be biased toward higher socioeconomic status. Finally, recall bias is another possibility due to the retrospective data collection. In spite of these limitations, this study had access to the largest available cohort of individuals with 3q29Del, which allowed us to gain initial insight into the psychosocial impacts of diagnosis.

### Practice Implications

Historically, non-genetics HCPs have not routinely ordered genetic testing as a diagnostic tool. Moving forward, genetic testing is now often ordered by primary care providers and non-genetics specialists. However, these ordering providers often consult and/or refer to genetics HCPs to interpret genetic testing results and to counsel the patient and their families.^31^ Thus, it will become even more important for non-genetics HCPs to become more familiar with identifying a need for genetic testing and ordering genetic testing appropriately in order to decrease the time to diagnosis. This shift will require an increase in genetics training and educational resources for non-genetics HCPs as well as interest and commitment from these HCPs.^31,32^

These results provide insight into the diagnostic experience of 3q29Del caregivers and help to highlight what caregivers need from HCPs at each time point in the diagnostic journey. During pre-test counseling, HCPs should provide appropriate anticipatory guidance for caregivers and explain testing limitations. Caregivers should also be informed of the possibility of finding a diagnosis that is not yet well described and may be associated with a high degree of variability. HCPs should be prepared to discuss and validate caregivers’ feelings surrounding the phenotypic variability, uncertainty, and association with psychiatric symptoms, and provide educational and support resources. Making these investments may strengthen the patient-doctor relationship and improve caregiver experiences. These results are likely similar to the experiences and feelings of caregivers of children with other variable genetic conditions. Thus, these recommendations may be applicable to additional patient populations.

### Research Recommendations

Studies are currently underway to better understand the phenotypic spectrum and long-term prognosis of 3q29Del.^5^ In the meantime, these data provide insights into the caregiver experience around the time of diagnosis of 3q29 deletion syndrome. Based on these findings, we recommend that HCPs (1) communicate the long-term risk for neuropsychiatric disease to caregivers, and (2) help families connect to resources for further information with other rare variant caregiver families. These relatively small changes may improve the diagnostic experience for caregivers, provide additional support for families, and promote access to early interventions.

## Data Availability

De-identified data will be made available upon request.

## Acknowledgements

We would like to thank the interviewees and their families for sharing their experiences. This study would not have been possible without their participation. We would also like thank Dr. Joseph Cubells, MD, PhD, Dr. Cecelia Bellcross, PhD, CGC, Kristen Cornell, MS, CGC, and Dr. Stephen Warren, PhD, FACMG for their help with this project. This study was supported by NIH Grants MH100917 and GM097331 and by the Emory University Treasure Your Exceptions Fund.

